## Supplementary material for "Combined multiplex panel test results are a poor estimate of disease prevalence without adjustment for test error": S1 Appendix

The rationale for this analysis is given in detail in the main paper, but in summary it is to investigate the properties of the result of a panel combination of multiple component tests. The component tests aim to detect individual subtypes of a condition which may appear independently of one another, but the presence of any subtype implies the presence of the index condition. Likewise, the index condition can be considered to be absent if none of the components are present. If we consider a single patient $k$ and a set of $N$ condition subtypes then the patient's condition status is given by:

$$I(A_{N,k}) = 1 - \prod_{n \in N} (1 - I(A_n, k))$$

Where $I$ is the indicator function and a value of one represents presence and zero is absence, and $A_n, k$ describes the actual status of the disease subtype $n$ in an individual $k$.

Such as scenario may occur when considering a single disease, that may be caused by multiple distinct pathogens which are tested for separately, and could, at least in theory, occur together.

#### 1 Definitions

Consider a set of patients $(K)$ who we are testing for disease. Their true disease status may be either positive or negative, we denote each individual true disease state with the binary value $A_k$. We then define the probability of disease over all patients as $P(A)$ which is the true prevalence of the disease $(prev)$. This quantity cannot necessarily be directly observed, and is usually something we wish to estimate - (N.b. $I$ is the indicator function where 1 is a positive and 0 a negative.)

$$prev = P(A) \approx \frac{\sum_{k \in K} I(A_k)}{|K|}$$

We assume all patients are tested for disease and there is a binary observation of a test result ($O_k$) for each $k$ of $K$ patients. The probability of observing a positive test result ($P(O)$), depends on disease status and test error, and is the apparent prevalence ($E(AP)$) which can be estimated using an observation of the test positivity ($\widehat{AP}$)

$$P(O) = E(AP) \approx \widehat{AP} = \frac{\sum_{k \in K} I(O_k)}{|K|}$$

Type 1 test errors are false positives, the rate of which may be defined as the probability of observing a positive test result given an actual negative case ($P(O|\neg A)$). The complement of this quantity is the true negative rate and is known as the specificity ($spec$). Type 2 test errors are false negatives and the rate of these is the probability of observing a negative test result given an actual positive case ($P(\neg O|A)$). The complement of this quantity is the true positive rate and is known as the sensitivity ($sens$).

$$\begin{aligned} spec &= TNR = P(\neg O|\neg A) = 1 - P(O|\neg A) = 1 - FPR \\ sens &= TPR = P(O|A) = 1 - P(\neg O|A) = 1 - FNR \end{aligned}$$

The likelihood that any given person is actual case positive and observed test positive is the true positives $TP$, and can be defined in terms of the sensitivity and prevalence ($sens$ and $prev$).

$$\begin{aligned} P(TN) &= P(\neg O|\neg A)P(\neg A) = spec \times (1 - prev) \\ P(FP) &= P(O|\neg A)P(\neg A) = (1 - spec) \times (1 - prev) \\ P(FN) &= P(\neg O|A)P(A) = (1 - sens) \times prev \\ P(TP) &= P(O|A)P(A) = sens \times prev \end{aligned} \tag{1}$$

#### 1.1 Rogan-Gladen derivation

An estimator for true prevalence presented by Rogan and Gladen [1] can be expressed in this notation from Eq (1) as:

$$\begin{aligned} P(O) &= P(TP) + P(FP) \\ P(O) &= P(O|A)P(A) + P(O|\neg A)P(\neg A) \\ P(O) &= P(O|A)P(A) + P(O|\neg A)(1 - P(A)) \\ P(O) &= P(O|A)P(A) + P(O|\neg A) - P(O|\neg A)P(A) \\ P(A) &= \frac{P(O) - P(O|\neg A)}{P(O|A) - P(O|\neg A)}. \end{aligned}$$

Or, expressed in more familiar terms, the apparent prevalence ($E(AP)$) from prevalence, sensitivity and specificity is described, which can be rearranged to get an estimate for true prevalence ($\widehat{prev}$) based on an observed test positive rate ($\widehat{AP}$), and sensitivity and specificity:

$$\begin{aligned} E(AP) &= sens \times prev + (1 - spec) \times (1 - prev) \\ \widehat{prev} &\approx \begin{cases} 0 & \widehat{AP} \leq (1 - spec) \\ \frac{\widehat{AP} + spec - 1}{sens + spec - 1} & (1 - spec) < \widehat{AP} < sens \\ 1 & sens \leq \widehat{AP} \end{cases} \end{aligned} \tag{2}$$

The estimator is only approximate because the use of an observed test positive rate ($\widehat{AP}$) as an estimate for the expected value of the apparent prevalence ($E(AP)$) becomes unreliable at extreme values. It also requires knowledge of test sensitivity and specificity.

An equivalence formulation for the Rogan-Gladen estimator using the complement of prevalence can be expressed as follows and will be referred to later:

$$
\begin{aligned}
P(\neg O) &= P(FN) + P(TN) \\
P(\neg O) &= P(\neg O|A)P(A) + P(\neg O|\neg A)P(\neg A) \\
P(\neg A) &= \frac{P(\neg O) - P(\neg O|A)}{P(\neg O|\neg A) - P(\neg O|A)} \\
P(\neg A) &= \frac{P(\neg O) - FNR}{TNR - FNR}.
\end{aligned} \tag{3}
$$

#### 2 Test panel combination

In the situation where a panel of independent tests looking for independently occurring features is combined, if any of the independent component test results are positive, we may infer the observed panel combination is positive. On the other hand only if all the test results are negative the panel may be considered as negative. If we again consider a single patient $k$ and a panel of component tests $N$, then the patient's combined result of the panel of tests is:

$$I(O_{N,k}) = 1 - \prod_{n \in N} (1 - I(O_n, k)).$$

Due to false positives and false negatives in the individual component tests however, the result of the observed combined panel will be inaccurate as compared to the actual index condition. If we consider an example panel consisting of two component tests, then the possible combination of results of a panel can be seen in table 1. The combination of imperfect test results result in an imperfect panel result. There are three things to note, all of which can be extended to scenarios involving more than two components. Firstly a panel result can only be considered a "true negative" (TN) result if all the component tests are themselves "true negative" (cyan). Secondly, the panel result is negative, if, and only if, all the component test results are negative, and unless all components are "true negatives", the negative panel result will be a "false negative" (yellow). Thirdly there is a class of outcomes when a panel will be positive, due to a combination of component false positives and false negatives, which is the correct panel result, but for the wrong reason (red). These results increase the effective true positive rate of the panel, over and above the true positive rates of the components.

Table 1:

| | ***test 2*** | ***pos*** | | ***neg*** | |
|---|---|---|---|---|---|
| ***test 1*** | | **TP** | **FP** | **FN** | **TN** |
| ***pos*** | **TP** | TP | TP | TP | TP |
| | **FP** | TP | FP | TP+ | FP |
| ***neg*** | **FN** | TP | TP+ | FN | FN |
| | **TN** | TP | FP | FN | TN |

#### 2.1 Actual prevalence of a condition as a collection of subtypes

Assuming independence of components, and complete knowledge of the subtypes, the probability of a patient having the supertype condition ($P(A_N)$) that results from any of the $N$ condition subtypes being present, is simply the complement of the probability that none of the condition subtypes is present:

$$
\begin{aligned}
prev_N &= 1 - \prod_{n \in N} (1 - prev_n), \\
1 - P(A_N) &= \prod_{n \in N} 1 - P(A_n), \\
P(\neg A_N) &= \prod_{n \in N} \frac{P(\neg O_n) - P(\neg O_n | A_n)}{P(\neg O_n | \neg A_n) - P(\neg O_n | A_n)}.
\end{aligned}
$$

#### 2.2 Observed test positivity of a panel as a collection of component tests

Assuming independence of components, the observed positivity of a panel ($P(O_N)$) that is composed of $N$ component tests is simply the complement of the probability that none of the component tests is positive. This is also the expected value of the apparent prevalence of a panel test result ($E(AP_N)$):

$$
\begin{aligned}
P(O_N) &= 1 - \prod_{n \in N} 1 - P(O_n) \\
&= 1 - \prod_{n \in N} P(\neg O_n) \\
&= 1 - \prod_{n \in N} \Big( P(\neg O_n | \neg A_n) P(\neg A_n) + P(\neg O_n | A_n) P(A_n) \Big), \\
E(AP_N) &= 1 - \prod_{n \in N} \Big( TNR_n \times (1 - prev_n) + FNR_n \times prev_n \Big) \\
&= 1 - \prod_{n \in N} \Big( spec_n \times (1 - prev_n) + (1 - sens_n) \times prev_n \Big).
\end{aligned}
$$

#### 2.3 Panel specificity: $TNR = 1 - FPR$

As we discussed above and in table 1, for a panel of $N$ tests, the panel result is a true negative, if and only if all the $N$ test results are also true negatives. From the definition of the probability of a given result being a true positive in (1), we can derive the true negative rate, i.e. the specificity:

$$
\begin{aligned}
P(TN_N) &= P(\neg O_N|\neg A_N)P(\neg A_N) \\
&= \prod_{n\in N} P(\neg O_n|\neg A_n)P(\neg A_n), \\
TNR_N &= P(\neg O_N|\neg A_N) \\
&= \prod_{n\in N} P(\neg O_n|\neg A_n), \\
spec_N &= \prod_{n\in N} spec_n.
\end{aligned} \tag{4}
$$

#### 2.4 Panel sensitivity: $TPR = 1 - FNR$

We determined above that if all observed component results are negative, the panel is also negative and that this is a false negative unless all component results are true negatives. This can also be derived from the definition of probability of the false negative result (1), using the following application of Bayes theorem:

$$
\begin{aligned}
P(\neg Y|X)P(X) &= P(X|\neg Y)P(\neg Y) \\
&= (1 - P(\neg X|\neg Y))P(\neg Y) \\
&= P(\neg Y) - P(\neg X|\neg Y)P(\neg Y) \\
&= P(\neg Y) - P(\neg Y|\neg X)P(\neg X).
\end{aligned}
$$

When this is applied to the definition of the probability that an observation is a false negative, we can derive an expression for the false negative rate, i.e. 1 - sensitivity, from (1):

$$
\begin{aligned}
P(FN_N) &= P(\neg O_N|A_N)P(A_N) \\
&= P(\neg O_N) - P(\neg O_N|\neg A_N)P(\neg A_N) \\
&= \prod_{n\in N} P(\neg O_n) - \prod_{n\in N} P(\neg O_n|\neg A_n)P(\neg A_n), \\
FNR_N &= P(\neg O_N|A_N) \\
&= \frac{P(FN_N)}{P(A_N)} \\
&= \frac{\prod_{n\in N} P(\neg O_n) - \prod_{n\in N} P(\neg O_n|\neg A_n)P(\neg A_n)}{1 - \prod_{n\in N} P(\neg A_n)}.
\end{aligned}
$$

From this and the Rogan-Gladen estimates in (3) we can eliminate $P(\neg O_n)$ terms, giving us a theoretical value for the sensitivity of the panel ($sens_N$) calculated using the prevalence of actual disease, if we know it, and component test sensitivity and specificity. This can be used to express the panel sensitivity in a Bayesian framework:

$$
\begin{aligned}
FNR_N &= \frac{\prod_{n\in N}\Big(P(\neg O_n|A_n)P(A_n) + P(\neg O_n|\neg A_n)P(\neg A_n)\Big) - \prod_{n\in N} P(\neg O_n|\neg A_n)P(\neg A_n)}{1 - \prod_{n\in N} P(\neg A_n),} \\
sens_N &= 1 - \frac{\prod_{n\in N}\Big((1 - sens_n) \times prev_n + spec_n \times (1 - prev_n)\Big) - \prod_{n\in N} spec_n \times (1 - prev_n)}{1 - \prod_{n\in N}(1 - prev_n)}.
\end{aligned} \tag{5}
$$

Alternatively using (3) we can eliminate $P(\neg A_n)$ terms, to express an estimate of panel sensitivity ($\widehat{sens_N}$) in terms of a set of observations of component test positive results ($\widehat{AP_n}$), and associated component test sensitivity and specificity:

$$
\begin{aligned}
FNR_N &= \frac{\prod_{n\in N} P(\neg O_n) - \prod_{n\in N} TNR_n \frac{P(\neg O_n)-FNR_n}{TNR_n-FNR_n}}{1-\prod_{n\in N}\frac{P(\neg O_n)-FNR_n}{TNR_n-FNR_n}},\\
\widehat{sens_N} &\approx 1 - \frac{\prod_{n\in N}(1-\widehat{AP_n}) - \prod_{n\in N} spec_n \times \frac{(1-\widehat{AP_n})-(1-sens_n)}{spec_n-(1-sens_n)}}{1-\prod_{n\in N}\frac{(1-\widehat{AP_n})-(1-sens_n)}{spec_n-(1-sens_n)}}\\
&\approx 1 - \frac{\prod_{n\in N}(1-\widehat{AP_n}) - \prod_{n\in N} spec_n \times \frac{sens_n-\widehat{AP_n}}{spec_n+sens_n-1}}{1-\prod_{n\in N}\frac{sens_n-\widehat{AP_n}}{spec_n+sens_n-1}}.
\end{aligned} \tag{6}
$$

This alternative expression is an estimator for panel specificity based on observed data, but as with the Rogan-Gladen estimate for true prevalence, it may become inaccurate when the observations of component test positivity is a poor proxy of expected apparent prevalence (i.e. outside of the range $(1-spec_n) < \widehat{AP_n} < sens_n$). However, in practice, this is less of an issue than the Rogan-Gladen estimator, and truncation is not required to produce reasonable estimates of combined sensitivity.

##### 2.5 Panel prevalence from data

These expressions allow us to create an estimator the "actual" prevalence of a condition using the observed test positivity of a panel of component tests by applying the Rogan-Gladen estimator. This can be calculated from raw component test data, and component test sensitivity and specificity:

$$
\begin{aligned}
\widehat{prev_N} &\approx \frac{\widehat{AP_N} + spec_N - 1}{\widehat{sens_N} + spec_N - 1}\\
&\approx \frac{\widehat{AP_N} + \prod_{n\in N} spec_n - 1}{\prod_{n\in N} spec_n - \frac{\prod_{n\in N}(1-\widehat{AP_n}) - \prod_{n\in N} spec_n \times \frac{sens_n-\widehat{AP_n}}{spec_n+sens_n-1}}{1-\prod_{n\in N}\frac{sens_n-\widehat{AP_n}}{spec_n+sens_n-1}}}\\
&\approx \frac{\prod_{n\in N} spec_n - (1-\widehat{AP_N})}{\prod_{n\in N} spec_n - \prod_{n\in N}(1-\widehat{AP_n})}\left(1-\prod_{n\in N}\frac{sens_n-\widehat{AP_n}}{spec_n+sens_n-1}\right).
\end{aligned} \tag{7}
$$

This expression is equivalent to the Rogan-Gladen estimate for panel tests and has a similar set of caveats and requirement for truncation of the prevalence estimate at high and low values of apparent prevalence.

#### 3 Characterisation

The combined prevalence, combined apparent prevalence, and combined specificity are a straightforward compound of their components, and behave in an unsurprising fashion.

In both theoretical (5) and estimator (6) forms of the combined sensitivity there is an interaction between component sensitivity, component specificity and component prevalence. This means that interpretation of the observed positivity of a panel test is not at all trivial, and unlike normal tests, the false negative rate is dependent on prevalence, with higher prevalence having lower false negative rate. There are a large number of free parameters and fully characterising the behaviour of this relationship is difficult so we restrict ourselves to some illustrative examples.

In figure 1, panel A, this relationship is demonstrated in an artificial scenario where we assume 10 identically prevalent diseases, and fix 10 component tests to all have a specificity of 0.975. The sensitivities of all components are varied together between 0.5 and 1 and the prevalence of all components varied between 0 and 0.25. The sensitivity of the combined test (from (5)) is always higher than that of the components, and this is increased with increasing prevalence. This is the result of more combined tests being correct for the wrong reason, as false positives in one test component balance out false negatives in another test component.

Secondly, in figure 1, panel B we explore how the combined sensitivity varies with varying sensitivities of the component tests. In this scenario all 10 components have the same prevalence (0.05) and all 10 component tests have the same specificity (0.975). In this case we set the sensitivity of the components to be either low (baseline - grey line) or high (black line) and vary the ratio of high versus low sensitivity tests, as we vary the baseline sensitivity between 0.5 and 1. In this artificial situation the combined sensitivity appears to be proportional to the average of the component sensitivity.

Thirdly in figure 1, panel C we explore the relationship between component test specificity and combined sensitivity, by fixing the prevalence of all components to 0.05. The sensitivities of all components are varied together between 0.5 and 1 and the specificity of all components varied between 0.75 and 1. In this scenario we see that the combined sensitivity is increased by decreasing component specificity, for the same reason as before, that decreasing the specificity of the components will result in more false positives to balance out any false negatives in another component.

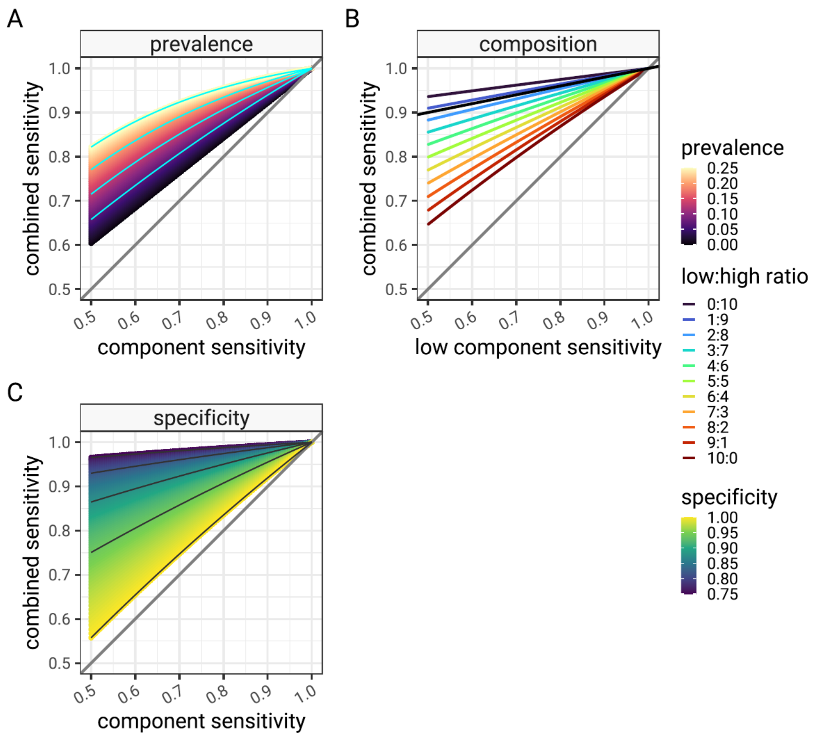


Figure 1: The relationship between component sensitivity and combined panel sensitivity for a panel of 10 tests. In the three subfigures A,B and C, the component tests are all kept the same, but one of disease prevalence, panel composition and component specificity are varied. In all cases the panel sensitivity is calculated using equation (6) for a range of component sensitivities. In this artificial scenario, where a given parameter is not being varied all components are set to have the same prevalence (0.05), test specificity (0.975) and test sensitivity is varied between 0.5 and 1.

### 4 Simulation

Whilst the characterisation is helpful to test the estimator in a wide range of scenarios, we resort to multiple simulations for further detailed characterisation. Following the model described in (8), we run 8 batches of 100 simulations. Each simulation consisting of between 2 and 9 components (depending on the batch) and each component in every simulation has a randomly assigned, but generally plausible, prevalence, sensitivity and specificity. Each component is given a random zero-inflated prevalence ($prev_n$) using a product of two Poisson distributed quantities, constrained so that the total prevalence matches a simulation parameter ($prev_{N_{sim}}$). 10000 cases are generated for each of the 800 simulations with "actual" subtype status depending on the component prevalence, and a test observation for each subtype depending on the "actual" subtype status, sensitivity and specificity. "Actual" combined disease status and observed test panel status are calculated for each of the 10000 cases. The model for each simulation is described below:

$$
\begin{aligned}
n &\in \{1, \dots, N_{sim}\}, \\
sens_n &\sim Beta(80, 20), \\
spec_n &\sim Beta(97.5, 2.5), \\
prev_N &\sim Beta(2, 10), \\
dist_n &\sim Poisson(5) \times Poisson(1), \\
prev_n &= \frac{dist_n}{\sum dist_n} \times prev_{N_{sim}}, \\
k &\in \{1, 2, \dots, 10000\}, \\
I(A_{n,k}) &\sim Bernoulli(prev_n), \\
I(O_{n,k}) &\sim Bernoulli((1 - spec_n) \times (1 - I(A_{n,k})) + sens_n \times I(A_{n,k})), \\
I(A_{N,k}) &= \prod_n I(A_{n,k}), \\
I(O_{N,k}) &= \prod_n I(O_{n,k}).
\end{aligned} \tag{8}
$$

For each simulation, the 10000 combined panel test status and the "actual" combined disease status are aggregated to provide a per-simulation estimate of panel sensitivity ($\widehat{sens_{N,sim}}$) and panel specificity ($\widehat{spec_{N,sim}}$), including confidence intervals, as defined in (10):

$$
\begin{aligned}
TP_N &= \sum_i I(A_{N,i}) \times I(O_{N,i}), \\
FP_N &= \sum_i I(\neg A_{N,i}) \times I(O_{N,i}), \\
FN_N &= \sum_i I(A_{N,i}) \times I(\neg O_{N,i}), \\
TN_N &= \sum_i I(\neg A_{N,i}) \times I(\neg O_{N,i}), \\
\widehat{spec_{N,sim}} &= \frac{TN_N}{FP_N + TN_N}, \\
\widehat{sens_{N,sim}} &= \frac{TP_N}{FN_N + TP_N}, \\
\widehat{AP_{N,sim}} &= \frac{1}{10000} \sum_i I(O_{N,i}).
\end{aligned} \tag{9}
$$

These per-simulation values are compared to the theoretical values of panel specificity ($spec_N$) (4), to the theoretical panel sensitivity derived from the simulation parameter "actual" prevalence ($sens_N$) obtained using (5) and to the estimated panel sensitivity ($\widehat{sens_N}$) derived from "observed" simulation component test apparent prevalence, obtained using (6). Lastly we can also estimate the "actual" prevalence of the overall disease by using the Rogan-Gladen estimator, observed panel test positives, predicted panel sensitivity and specificity. This can be directly compared to the parameterised simulation panel prevalence:

$$
\widehat{prev_{N,sim}} = \frac{\widehat{AP_{N,sim}} + spec_N - 1}{\widehat{sens_N} + spec_N - 1}. \tag{10}
$$

The primary results of the simulation are shown in figure 2. Panel A shows the relationship between the prevalence of each component disease as a provided simulation parameter and we see the effect of false positives in low prevalence, biasing the apparent prevalence high with respect to the actual value. In panel B, the components are adjusted using the Rogan-Gladen estimator, using simulation parameters for sensitivity and specificity correcting the bias.

Panel C shows the relationship between the apparent prevalence of the combined panel result compared to the actual prevalence of the combined disease as a provided simulation parameter. The scale of the error can be quite large as the panel prevalence is compounding multiple false positive errors from each component, depending on the sensitivity and specificity of the components. The scale of the error will depend on the number of components in the panel and the sensitivities and specificities of the components as described above. In Panel D, the Rogan-Gladen correction is applied, using the panel apparent prevalence, and the estimates of panel sensitivity and specificity from this analysis (equations (6) and (4)). The correction is effective in restoring an accurate estimate of the actual prevalence.

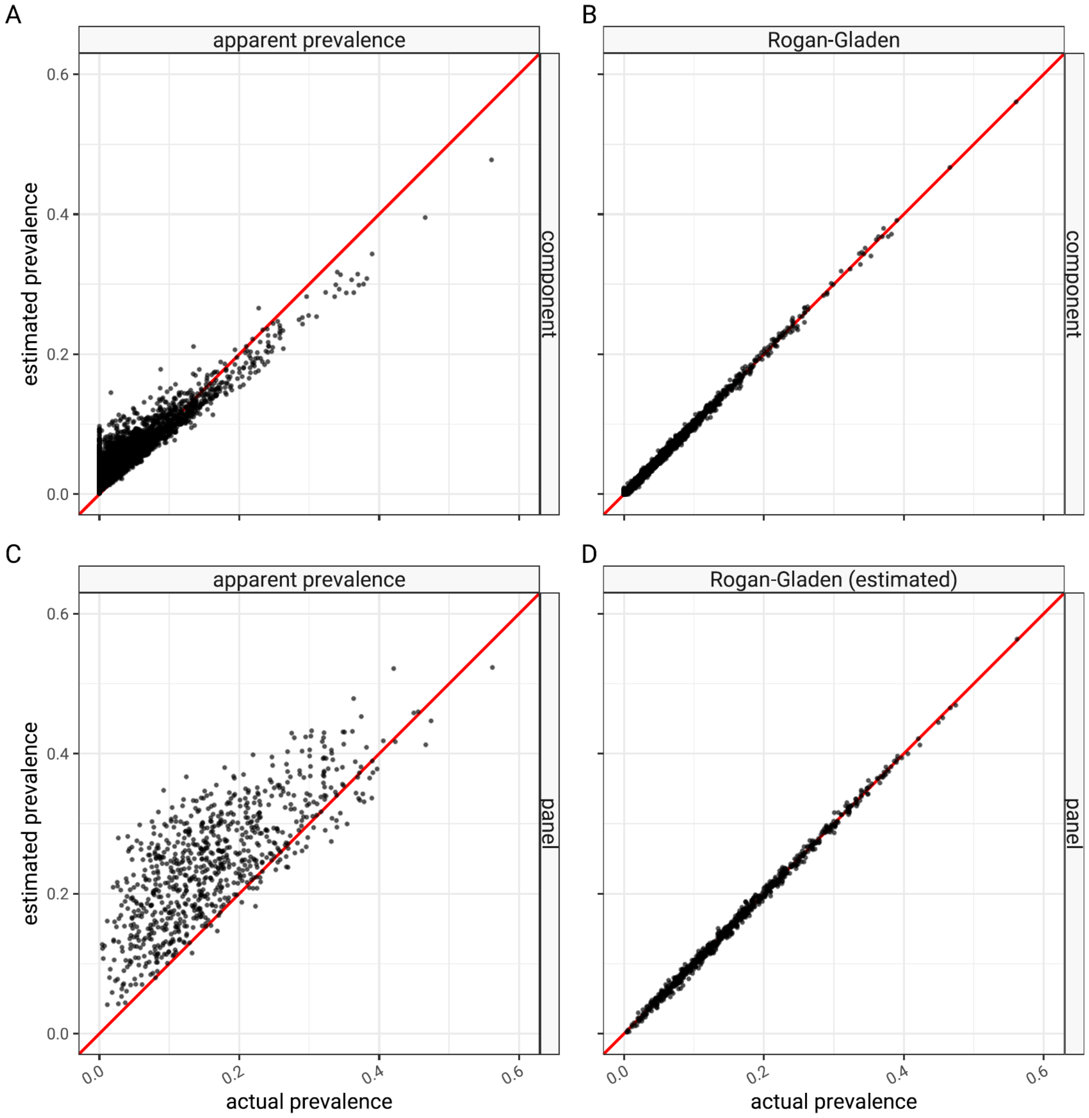


Figure 2: Apparent prevalence (uncorrected estimate) and Rogan-Gladen (corrected estimate) as compared to simulation prevalence. Panels A and B show results each of the 4400 single component tests in the 800 simulations (various component test sensitivities, specificities). Panels C and D show the results of the 800 combined panels (2-9 components per panel, various component distributions).

In panel A, figure 3, we also see a close relationship between the theoretical panel specificity and panel specificity estimated from each simulation scenario. As specificity is estimated from test negative cases the confidence intervals of the simulation estimates are quite narrow.

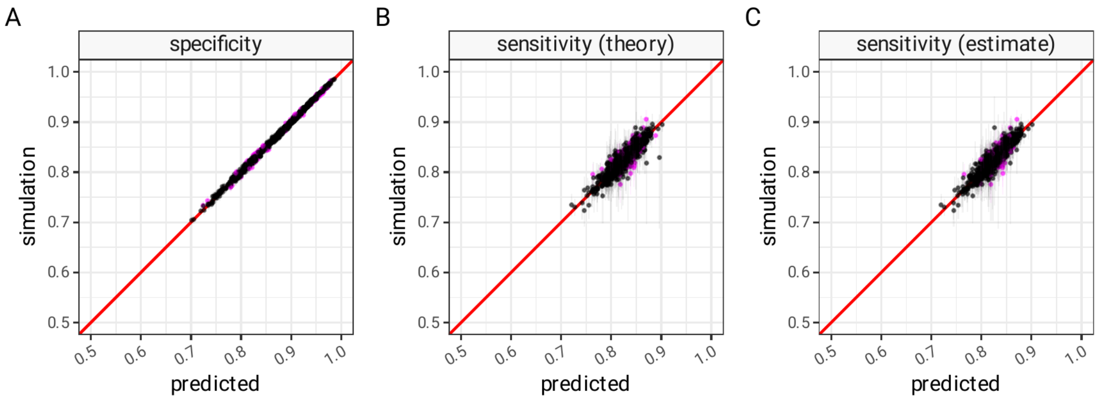


Figure 3: Panel sensitivity predicted using (4) (subfigure A) and specificity predicted using equations (5) (subfigure B) and equation (6) - (subfigure C) as compared to panel sensitivity and specificity values derived from the combined test positive and actual positives from simulated component data.

Panel B shows a comparison between predicted value of panel sensitivity (using (5) in this formal analysis) and the observed panel sensitivity from simulation. Panel C shows panel sensitivity predicted from component test positive rates that would be observed in a real setting ((6) in this analysis). In both cases there is a reasonable agreement between predicted and simulated values.

In both types of sensitivity estimate there is divergence between the predicted panel sensitivity and the observed simulation panel sensitivity. This is seen to be a worse in scenarios with low prevalence, as expected (figure 4). There is no clear trend in the error in scenarios with different sensitivities or in scenarios with different numbers of components. Errors in all cases are symmetrical suggesting the sensitivity predictions are unbiased.

If the theoretical values of sensitivity are accurate then it should lie within binomial confidence limits of the simulation estimates 95% of the time. Disagreement occurs when the theoretical value is outside the confidence interval. In figure 5 we ses the rates of disagreement as a function of panel prevalence, specificity and number of components. This demonstrates that in the areas of simulation parameter space where we have sufficient coverage, disagreement rates between simulated and theoretical values are close to the 0.05 value resulting from the 95% confidence limits in the simulation. There is no clear trend.

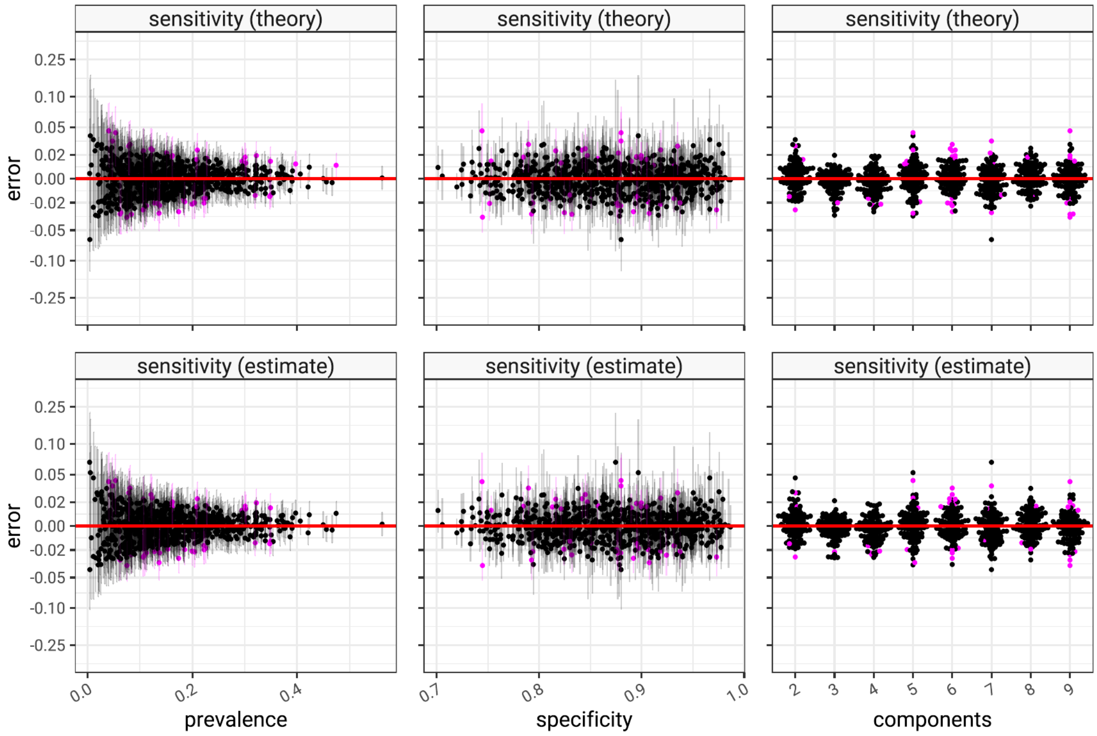


Figure 4: Error between panel sensitivity estimates predicted using equations (5) (top row) and equation (6) (bottom row) and observed simulation panel sensitivity. Error size is plotted as a function of simulation panel prevalence (first row), panel specificity (second row) and number of panel components (third row). Error magnitude depends on the quality of the estimator and the inherent error from using simulated data, which is higher when prevalence is low.

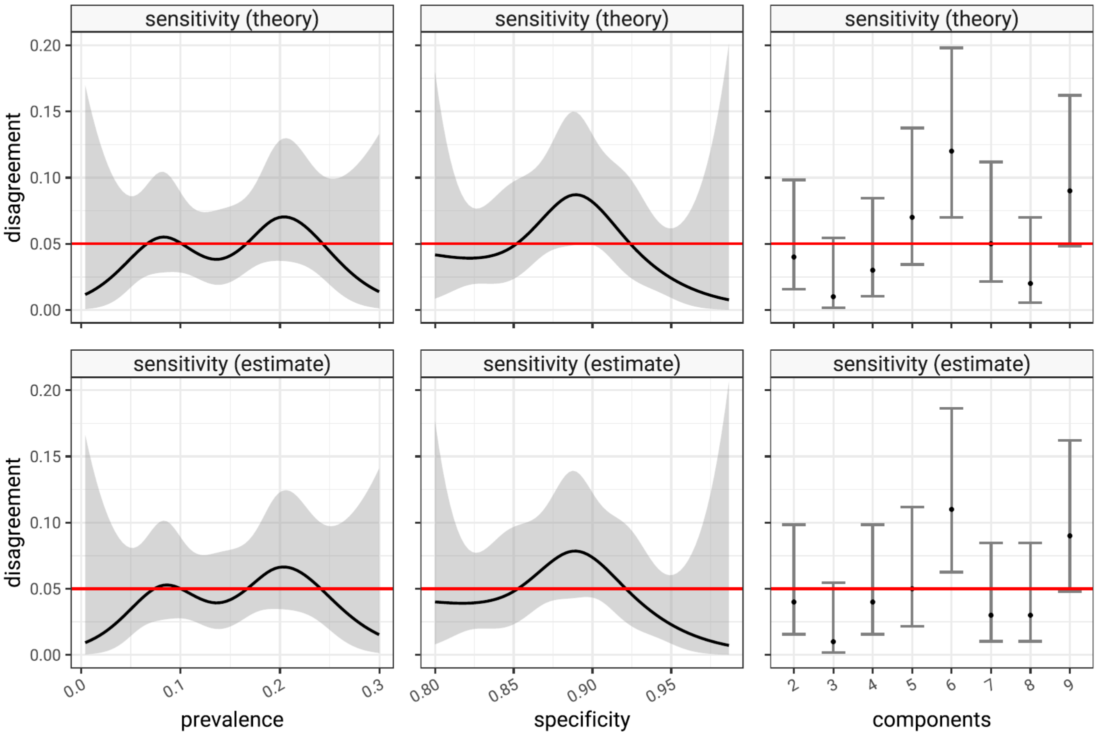


Figure 5: Percentage disagreement between panel sensitivity estimates predicted using equations (5) (top row) and equation (6) (bottom row) versus observed simulation panel sensitivity. Disagreement accounts for uncertainty introduced by simulation by comparing the binomial confidence intervals of simulation panel sensitivity estimates with sensitivity predictions. 95% confidence limits imply disagreement should be at the level of 5%. Disagreement rates are plotted as a function of simulation panel prevalence (first row), panel specificity (second row) and number of panel components (third row).

### 5 Summary

In this formal analysis we describe the relationship between panel specificity and sensitivity, and the component test sensitivities and specificities that comprise the panel. We demonstrate that panel specificity is the product of component specificities, and we demonstrate that panel sensitivity has a dependency on component sensitivities, component specificities and the prevalence of the subtypes of disease that are being tested for.

The implication of this is that when interpreting panel test results, we must take into account the finding that component false positive rates are compounded up in the panel result, and that false negative rates within a panel are not simple to predict and, unlike the usual concept of sensitivity, depend on the prevalence of disease in the sampled population.

We have derived expressions that can estimate panel sensitivity and specificity and taken together with the panel test positivity, we have demonstrated that a good central estimate of prevalence can be recovered from a simulation.

Propagation of uncertainty in these estimates is not trivial and this is addressed in Supplementary Appendix 2.
