## Supplementary material for "Combined multiplex panel test results are a poor estimate of disease prevalence without adjustment for test error": S2 Appendix

#### 1 Introduction

The problem of estimating expected prevalence or apparent prevalence (AP) from observed positive test results with imperfect tests is well known; Rogan and Gladen [7] described an adjustment to apparent prevalence to give an unbiased point estimate of true prevalence. In a set of patients $K$, the expected number of positive test results, or apparent prevalence, $E(AP)$, is a function of prevalence, test sensitivity ($sens$) and test specificity ($spec$). A single observation of apparent prevalence $\widehat{AP}$, is the rate of the positive test results per patient, $I_k$, and this can be used to estimate true prevalence ($prev$). When prevalence is low, apparent prevalence is an over-estimate due to false positives, and when high an underestimate due to false negatives. There is a critical value of prevalence ($prev_{crit}$) at which false positives and false negatives exactly balance and apparent prevalence is equal to true prevalence:

$$
\begin{aligned}
E(AP) &= prev \times sens + (1 - prev) \times (1 - spec), \\
\widehat{AP} &= \frac{1}{|K|} \sum_{k \in K} I_k, \\
prev &\approx \begin{cases} 0 & \widehat{AP} \leq (1 - spec) \\ \frac{\widehat{AP} + spec - 1}{sens + spec - 1} & (1 - spec) < \widehat{AP} < sens \\ 1 & sens \leq \widehat{AP_N}, \end{cases} \\
prev_{crit} &= \frac{(1 - spec)}{(2 - spec - sens)}.
\end{aligned}
$$

In Fig 1 these relationships are plotted for a hypothetical test with known sensitivity (80%) and specificity (95%). The density demonstrates sampling error of test positives in 100 observations (in the y-direction). At a true prevalence of 0.05, we expect to see about 9 test positives in 100 observations (blue curve left hand panel). However a single observation of test positivity may have arisen from a wide range of possible values of true prevalence (x-direction), and in this case 9 positive observations may be quite feasibly due to a true prevalence between 0 and about 0.15 (blue curve bottom panel). The test positives are not binomially distributed and binomial confidence intervals (blue bar in bottom panel) are biased with direction depending on prevalence. It has been proven that the variance of true prevalence is always larger than the variance of apparent prevalence [6, 7] and this is seen by the larger width than height of the probability density in Fig 1.

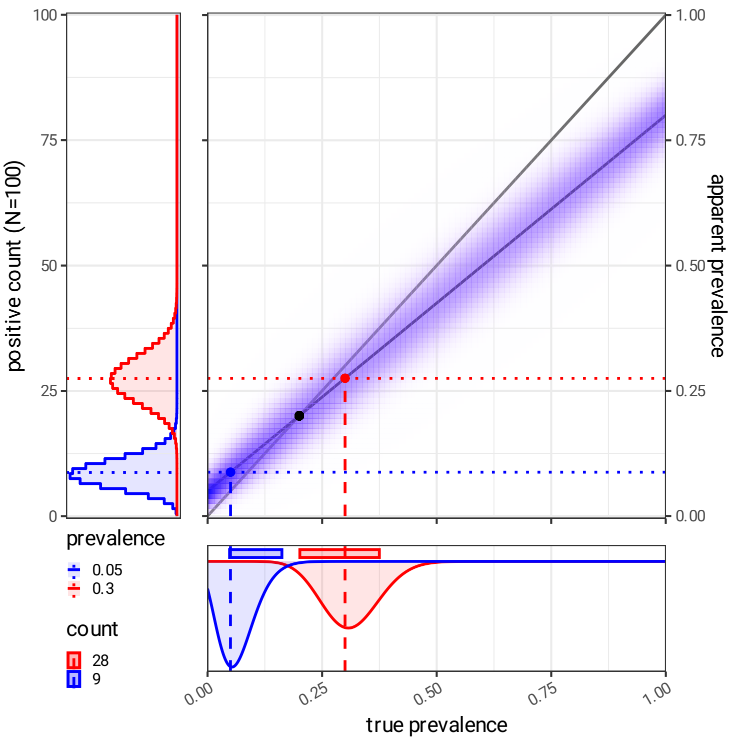


Figure 1: The relationship between true prevalence and expected test positivity for a test with specificity of 95%, and sensitivity of 80%. Shading represents the probability of observing a specific test positivity rate in 100 samples, which is also shown in the left panel for the examples of true prevalence of 0.05 (blue) and 0.3 (red). In this figure the exact sensitivity and specificity are assumed.

The uncertainty in true prevalence, from an observation of apparent prevalence, is also dependent on uncertainty in sensitivity and specificity. This was quantified by Lang and Reiczigel (2014) [3,6] in a frequentist framework and Gelman (2020) and Diggle (2011) [2–4] in a Bayesian one, to estimate confidence intervals of the true prevalence given uncertainty of both test sensitivity and specificity.

In this supplementary document, we extend these methods to the situation of multiplex testing where the uncertainty in sensitivity and specificity may apply to multiple components of a single test and where resulting test error is compounded.

### 2 Methods

#### 2.1 Simulation

We previously described a distribution of serotypes in invasive pneumococcal disease (IPD) [5]. To test the methods described here we used the IPD distribution and simulated a set of synthetic patients with known rates of disease super-types and subtypes, with a single example as shown in Fig 2. These synthetic patients were assigned component multiplex test results (in this case representing individual pneumococcal serotypes), assuming specific values of sensitivity and specificity. The simulated test results of the individual serotypes were then aggregated into four panels: a PCV7 group, a PCV13 group, a PCV15 group and a PCV20 group. The simulation was repeated for a range of defined prevalence levels, from 2.5% to 20%. In default scenarios component sensitivity was set at 80% and specificity kept at 99.75%, or varied between 60%,75% and 90% (sensitivity) and between 99.75% and 90% (specificity).

In the formal description of the simulation that follows, $N$ describes the 4 PCV panels, $n$ each individual component serotype. The simulation includes $k$ synthetic patients (1000 was used in all cases), and their simulated actual pneumococcal serotype status is represented by $A_{n,k}$. The simulation of observed test result taking into account error is $O_{n,k}$. Each serotype has a frequency, ($freq_n$), which is scaled by an empirically determined factor ($scale$) to make sure the simulation has the desired prevalence of PCV20 ($prev_{PCV20}$):

$$
\begin{aligned}
N &\in \{PCV7, PCV13, PCV15, PCV20\},\\
PCV7 &\in \{4, 6B, 9V, 14, 18C, 19F, 23F\},\\
PCV13 &\in \{1, 3, 5, 6A, 7F, 19A\} \cup PCV7,\\
PCV15 &\in \{22F, 33F\} \cup PCV13,\\
PCV20 &\in \{8, 10A, 11A, 12F, 15B\} \cup PCV15,\\
prev_{PCV20} &\in \{2.5\%, 5\% \dots 20\%\},\\
prev_n &= \frac{scale \times freq_n}{\sum freq_n},\\
prev_N &= 1 - \prod_{n \in N} 1 - prev_n,\\
sens_n &\in \{80\%, 60\%, 75\%, 90\%\},\\
spec_n &\in \{99.75\%, 90\%\},\\
A_{n,k} &\sim Bernoulli(prev_n),\\
O_{n,k} &\sim Bernoulli(A_{n,k} \times sens_n + (1 - A_{n,k}) \times (1 - spec_n)),\\
A_{N,k} &= 1 - \prod_{n \in N} 1 - A_{n,k},\\
O_{N,k} &= 1 - \prod_{n \in N} 1 - O_{n,k},\\
\widehat{AP} &= \frac{1}{|K|} \sum_{k \in K} I(O_k).
\end{aligned}
$$

Methods for propagating uncertainty described below (Bayesian, Lang-Reiczigel, and resampled Rogan-Gladen) were tested using matching, and mismatching prior assumptions for component sensitivity and specificity including uncertainty, and the resulting predictions for true prevalence compared to the simulated true prevalence. As there are a large number of free parameters, and some of the methods are computationally expensive, only a restricted number of representative scenarios were tested.

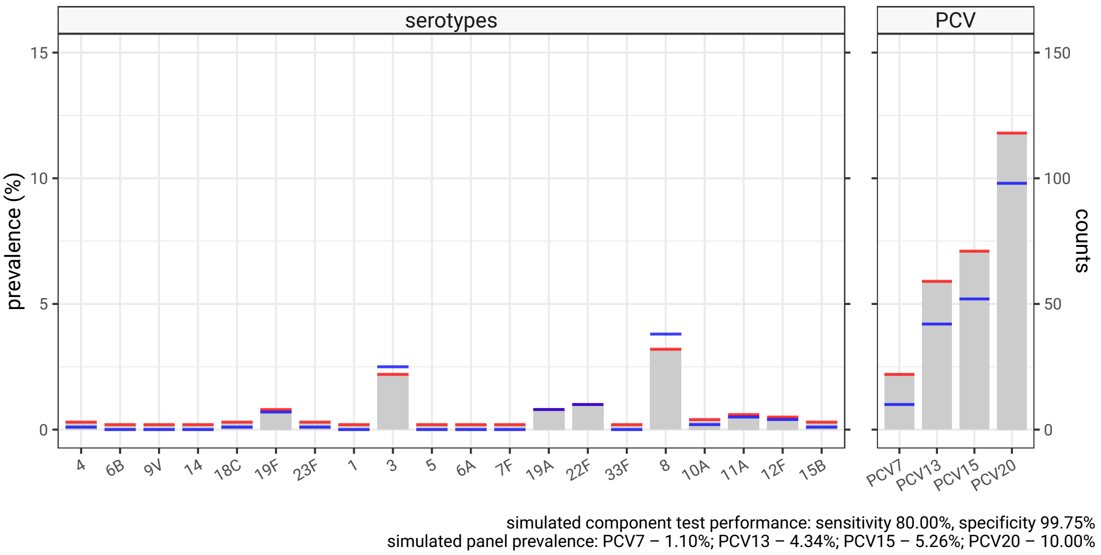


Figure 2: The relative frequency of the 20 pneumococcal serotypes contained in PCV20, and identified in invasive pneumococcal disease cases within the last 2 years, were converted to a distribution of 20 subtypes to give an overall PCV20 pneumococcal prevalence of 10% (blue lines). Test positive samples were created assuming each serotype test had a sensitivity of 80% and a specificity of 99.75% (red lines) showing a mix of test positivity as an underestimate of true prevalence (serotypes 3 and 8) and as an overestimate (the remainder). The simulated test result of the individual serotypes were aggregated into a PCV7 group (consisting of serotypes 4, 6B, 9V, 14, 18C, 19F, 23F), a PCV13 group (PCV7 groups plus 1, 3, 5, 6A, 7F, 19A), a PCV15 group (PCV13 plus 22F and 33F) and a PCV20 group (all serotypes). In the right subfigure, combined test positivity for the groups (apparent prevalence - red lines) all overestimate true prevalence (blue line) for this scenario.

#### 2.2 Rogan-Gladen estimator with resampling

As illustrated in Fig 1, there are three sources of uncertainty in estimates of true prevalence; there is uncertainty in test sensitivity, test specificity and observed test positivity. In the panel test there are three per component test which are combined in a non-linear fashion. A simple empirical method involves creating a set of randomly sampled component test sensitivity, specificity and apparent prevalence ($J$). Uncertainty in sensitivity is expressed as a Beta distributed quantity defined in terms of a disease positive control group, which consists of true positives ($TP_{disease^+}$) and false negatives ($FN_{disease^+}$). Specificity on the other hand is defined in terms of a disease negative control group, consisting of true negatives ($TN_{disease^-}$) and false positives ($FP_{disease^-}$). Apparent prevalence is assumed to originate from a binomial sample of size $k$ representing the number of patients tested.

With the sampled set $J$ we can directly apply the Rogan-Gladen estimator to derive a set of estimates of component prevalence including uncertainty. The empirical quantiles of these can be used as estimators of component prevalence.

Using the set $J$ we also calculate panel test sensitivity and specificity using the methods described in supplementary S1, and use these, and panel apparent prevalence in a Rogan-Gladen estimator [7] to create a set of estimates for true prevalence of the panel. The empirical quantiles and mean of this are taken as estimators for the true panel prevalence including uncertainty.

$$
\begin{aligned}
sens_{n,j} &\sim Beta(TP_{disease^{+},n}, FN_{disease^{+},n}),\\
spec_{n,j} &\sim Beta(TN_{disease^{-},n}, FP_{disease^{-},n}),\\
AP_{n,j} &\sim \frac{1}{k} Binomial(k, \widehat{AP_n}),\\
spec_{N,j} &= \prod_{n \in N} spec_{n,j},\\
sens_{N,j} &\approx 1 - \frac{\prod_{n \in N}(1 - AP_{n,j}) - \prod_{n \in N} spec_{n,j} \times \frac{sens_{n,j} - AP_{n,j}}{spec_{n,j} + sens_{n,j} - 1}}{1 - \prod_{n \in N} \frac{sens_{n,j} - AP_{n,j}}{spec_{n,j} + sens_{n,} - 1}},\\
prev_{N,j} &= \begin{cases} 0 & \widehat{AP_N} \leq (1 - spec_{N,j}) \\ \frac{\widehat{AP_N} + spec_{N,j} - 1}{sens_{N,j} + spec_{N,j} - 1} & (1 - spec_{N,j}) < \widehat{AP_N} < sens_{N,j} \\ 1 & sens_{N,j} \leq \widehat{AP_N} \end{cases},\\
\overline{prev_N} &= \frac{1}{|J|} \sum_{j \in J} prev_{N,i}.
\end{aligned}
$$

Confidence intervals are determined from the empirical quantiles of $prev_{N,j}$, $Q_{emp}(prev_{N,j}; z_{crit})$ and $Q_{emp}(prev_{N,j}; 1 - z_{crit})$.

#### 2.3 Lang-Reiczigel estimator

The Lang-Reiczigel estimator [6] includes uncertainty in test sensitivity and specificity which can be directly used to estimate the uncertainty in prevalence of component tests. There may be uncertainty in published data on panel test sensitivity and specificity, which can be adopted to use directly with a Lang-Reiczigel estimator. However this is not often available. In our example here, we combine component tests in multiple ways to generate 4 different PCV groups, each of which act as having their own sensitivity and specificity. Such combinations are unlikely to have published sensitivity and specificity. It should also be considered that panel test specificity is a function of component prevalence so is not generalisable between different sampled populations.

To deal with this, we again express uncertainty in sensitivity and specificity as Beta distributions and proceed to generate a set of samples as before. From these we derive a set of panel test specificity and sensitivity estimates as before:

$$
\begin{aligned}
sens_{n,j} &\sim Beta(TP_{disease^{+},n}, FN_{disease^{+},n}),\\
spec_{n,j} &\sim Beta(TN_{disease^{-},n}, FP_{disease^{-},n}),\\
AP_{n,j} &\sim \frac{1}{k} Binomial(k, \widehat{AP_n}),\\
spec_{N,j} &= \prod_{n \in N} spec_{n,j},\\
sens_{N,j} &\approx 1 - \frac{\prod_{n \in N}(1 - AP_{n,j}) - \prod_{n \in N} spec_{n,j} \times \frac{sens_{n,j} - AP_{n,j}}{spec_{n,j} + sens_{n,j} - 1}}{1 - \prod_{n \in N} \frac{sens_{n,j} - AP_{n,j}}{spec_{n,j} + sens_{n,j} - 1}}.
\end{aligned}
$$

We then calculate the parameters required for the Lang-Reiczigel method [6], which are expressed as central estimates of the panel sensitivity ($\widehat{Se}$) and specificity ($\widehat{Sp}$), and respective sample sizes (Beta distribution concentration parameters - $n_{Se}$ and $n_{Sp}$). We determine these from the set of panel test specificity and sensitivity estimates obtained above, by assuming matching the moments of their empirical distributions to that of a Beta distribution:

$$\widehat{Se} = \frac{1}{|J|}\sum sens_{N,j},$$
$$n_{Se} = \frac{\widehat{Se}(1-\widehat{Se})}{\frac{1}{|J|}\sum(sens_{N,j}-\widehat{Se})^2} - 1,$$
$$\widehat{Sp} = \frac{1}{|J|}\sum spec_{N,j},$$
$$n_{Sp} = \frac{\widehat{Sp}(1-\widehat{Sp})}{\frac{1}{|J|}\sum(spec_{N,j}-\widehat{Sp})^2} - 1.$$

These parameterised uncertain panel test sensitivity and specificity estimates are applied to Lang-Reiczigel equations (4 and 11-19) [6], to generate central estimates (which are calculated using the Rogan-Gladen formula) and confidence limits. Their method is not replicated here.

As the empirical distribution of panel test sensitivity and specificity is approximated by a Beta distribution we may expect additional uncertainty in this method, compared to resampling, however the central estimates are both generated using Rogan-Gladen methods so should be similar.

#### 2.4 Bayesian model

Both the methods described above both suffer from the same weakness in that at low (and high) prevalence, the Rogan-Gladen estimator must be truncated. This could lead to undesirable effects at low prevalence. In certain real life scenarios we do not have estimates of both sensitivity both specificity information at the individual component level, but rather at the level of the combined panel. In a Bayesian framework we can incorporate prior assumptions about the sensitivity and specificity with specific control group test results, at both component and panel levels to inform estimates of true prevalence.

In this model we are trying to estimate the true prevalence of $K_{sample}$ subjects tested with a panel with $n$ component subtypes. We assume informed priors for sensitivity and specificity expressed as a logistic transform of normal distributions with parameters $\mu$ and $\sigma$ (i.e. $P(\mathcal{N}(\mu,\sigma^2))$ where P is the standard logistic function), and a set of disease negative controls ($K_{disease^-}$) for specificity, and disease positive controls ($K_{disease^+}$) for sensitivity. To make this model identifiable some weak prior information is also required about the true prevalence of the components. We also require the component prevalence to follow a logistic normal distribution with parameters $\mu$ and $\sigma$. Based on the work of Gelman et al. [4], we propose the following model to describe the result of an individual's test result for a single component ($I(O_{n,k_{sample}})$ - with $I$ as an indicator of test positivity), using the two hyperparameters $\mu$ and $\sigma$ for the sensitivity, specificity, and prevalence of each component tests, and data $TP_{disease^+,n}$, $TN_{disease^-,n}$, and $I(O_{n,k_{sample}})$ describing the observed control group results and the sample under investigation repectively:

$$
\begin{aligned}
sens_n &\sim P(\mathcal{N}(\mu_{disease^+,n}, \sigma^2_{disease^+,n})),\\
spec_n &\sim P(\mathcal{N}(\mu_{disease^-,n}, \sigma^2_{disease^-,n})),\\
TP_{disease^+,n} &\sim Binomial(|K_{disease^+,n}|, sens_n),\\
TN_{disease^-,n} &\sim Binomial(|K_{disease^-,n}|, spec_n),\\
prev_n &\sim P(\mathcal{N}(\mu_{sample,n}, \sigma^2_{sample,n})),\\
AP_n &= prev_n \times sens_n + (1 - spec_n) \times (1 - prev_n),\\
\widehat{AP}_n &= \sum_{K_{sample}} I(O_{n,k_{sample}}),\\
\widehat{AP}_n &\sim Binomial(|K_{sample}|, AP_n).
\end{aligned}
$$

With the relationships determined in supplementary information S1 we can determine panel sensitivity and specificity, and aggregate component test results to panel results ($I(O_{N,k_{sample}})$) per test subject and hence observed test positivity in all subjects ($\widehat{AP_N}$):

$$
\begin{aligned}
prev_N &= 1 - \prod_{n \in N}(1 - prev_n),\\
spec_N &= \prod_{n \in N} spec_n,\\
sens_N &= 1 - \frac{\prod_{n \in N}\Big((1 - sens_n)prev_n + spec_n(1 - prev_n)\Big) - \prod_{n \in N} spec_n(1 - prev_n)}{1 - \prod_{n \in N}(1 - prev_n)},\\
I(O_{N,k_{sample}}) &= 1 - \prod_{n \in N} 1 - I(O_{n,k_{sample}}),\\
\widehat{AP_N} &= \sum_{K_{sample}} I(O_{N,k_{sample}}).
\end{aligned}
$$

From this we can also describe the observed panel apparent prevalence, and, if available, integrating any prior information we have about panel test sensitivity and specificity expressed as logistic normal distributions with hyperparameters $\mu$ and $\sigma$ as for the individual components:

$$
\begin{aligned}
sens_N &\sim P(\mathcal{N}(\mu_{disease^+,N}, \sigma^2_{disease^+,N})),\\
spec_N &\sim P(\mathcal{N}(\mu_{disease^-,N}, \sigma^2_{disease^-,N})),\\
AP_N &= prev_N \times sens_N + (1 - spec_N) \times (1 - prev_N),\\
\widehat{AP_N} &\sim Binomial(|K_{sample}|, AP_N).
\end{aligned}
$$

Maximising the log-likelihood of the combined model allows for simultaneous estimation of component and panel prevalence, and posterior estimates of component and panel sensitivity and specificity.

Choice of priors in this model depends on the nature of the control group data available. In all cases, component prevalence priors should only be weakly informative as it is a key output of the model. In this implementation, the $\mu$ value is automatically set to the Rogan-Gladen estimate and the $\sigma$ value set to a large value. If good control group data is available for control group specificity and sensitivity then prior estimates of these quantities can be weakly informative, and sensible default values encompassing a wide range of possibilities can be used. In this implementation we used $\mu = 2.217$, $\sigma = 2.202$, to give a wide range of sensitivity of 10.9% — 99.9% and a narrower $\mu = 18.47$, $\sigma = 8.966$, to give a range of specificites of 71% to 100%. A scenario encountered in real life is for reasonable amounts of control group data to be available for component specificity but very limited information for component sensitivity. In this case, we may have some additional information from studies of the performance of the panel test, which can be incorporated as a prior for panel sensitivity (further details are included in the documentation of the R package [1]).

### 3 Results

From Fig 1 we expect overestimation at low prevalence and underestimation at high prevalence. For the components this underestimation is clearly seen (Fig 3 left subfigure). Correction takes place and all methods produce estimates of true prevalence that are very close to the true value (blue line). For panel test results (Fig 3 right subfigure) the underestimation is much clearer as the panel specificity is lower that that of the individual components. Again, all correction methods result in an adjusted estimate closer to the true panel prevalence.

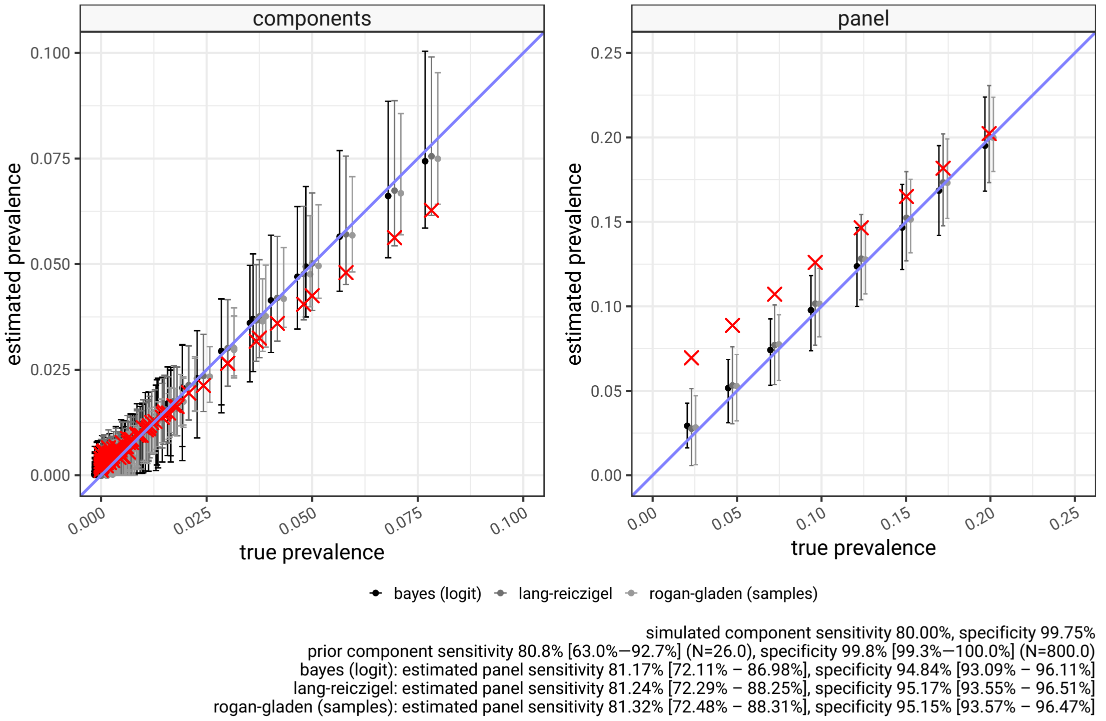


Figure 3: Apparent and adjusted prevalence estimates for component serotypes (right) and PCV20 panel (left) in the IPD simulation, at 8 different pre-set levels of overall prevalence. The red crosses show apparent prevalence, and the estimates of adjusted prevalence with associated uncertainty are shown in shades of grey, by methodology, including Bayesian, Rogan-Gladen or Lang-Reiczigel. The sensitivity and specificity parameters this simulation is based on are shown in the right margins and the prior estimates of component sensitivity and specificity that are used in the individual methods are shown in the top margins of each plot. In this example the priors are set to be equal to the simulation parameters but with uncertainty (displayed as 95% confidence intervals). All component tests are assumed to have the same sensitivity and specificity.

In Fig 4, left panels (A,C,E), the scenario is repeated for a range of different component test sensitivities. As expected, in the components the degree of underestimation at higher prevalence is related to sensitivity, but all three methods of correction behave similarly. Lower sensitivity levels result in larger confidence bounds. In the right panels (B,D,E) the panel adjusted prevalence is again close to the expected value which is contained in the confidence intervals.

In the results thus far, the adjustment has been done using prior estimates of sensitivity and specificity that are the same as those employed in the simulation. If however our prior assumptions around sensitivity are too high compared to simulation, the compensation will tend to push corrected true prevalence estimates too low (subfigure A in Fig 5). Conversely, if prior sensitivity is an overestimate, corrected true prevalence estimates will be too high (Fig 5 subfigure B). The accuracy of adjustment is more heavily influenced by prior assumptions of test specificity. Assuming too low a specificity causes frequentist methods to interpret all positives as false positives and in the majority of situations collapse estimates of true prevalence to zero (Fig 5, subfigure C), Bayesian approaches are better at compensating in this scenario, as the implausible combination of low test positives and low specificity is excluded. In the situation where specificity is assumed to be much higher than it is, the considerable false positive rate is misinterpreted as true positives and all methods fail to adjust (Fig 5, subfigure D). These mis-specifications are very large and in reality some degree of informed prior for sensitivity is required but more importantly for specificity.

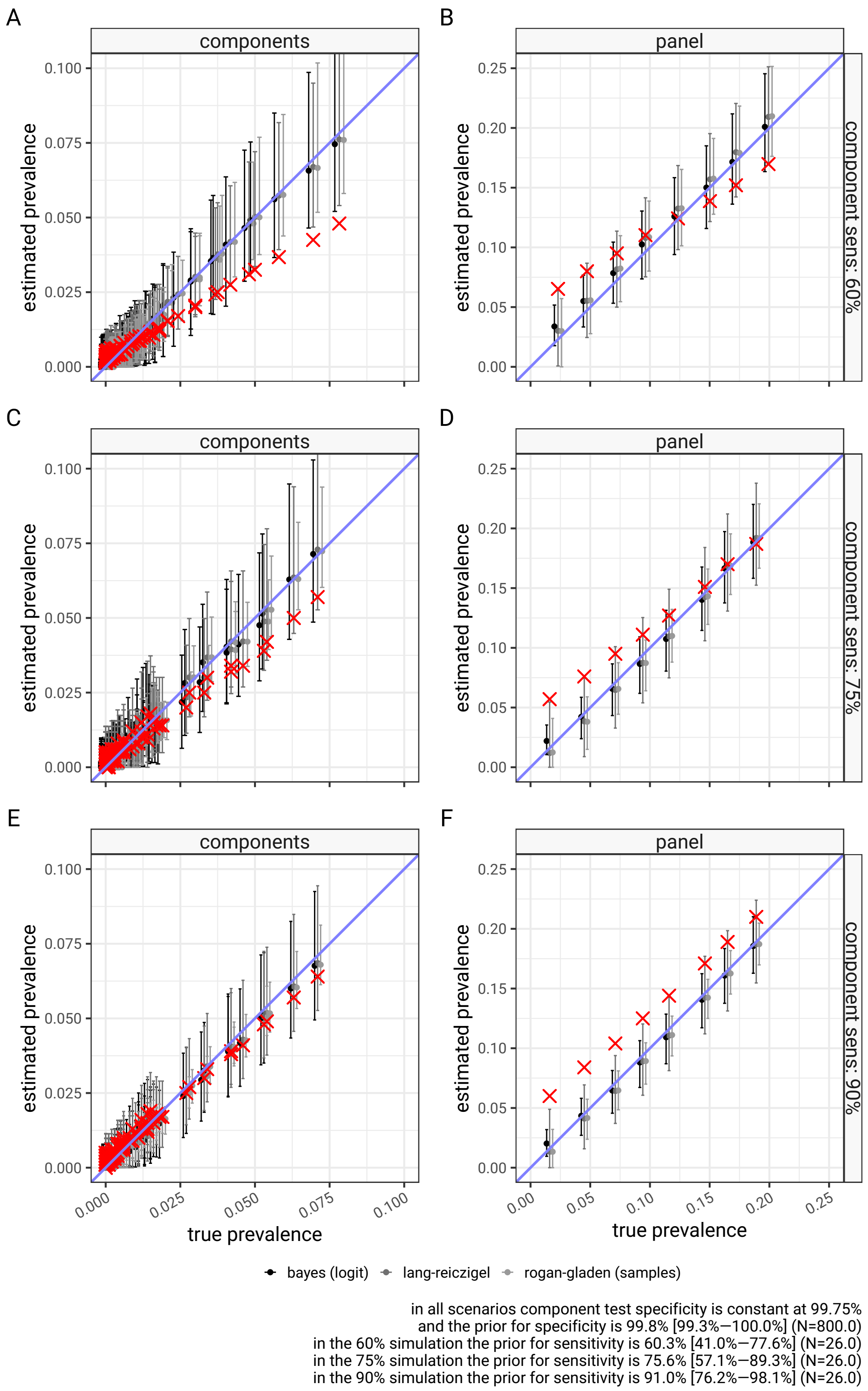


Figure 4: Apparent and adjusted prevalence of components and panels in a range of scenarios where tests sensitivity is varied. The red crosses show apparent prevalence, and the estimates of adjusted prevalence with associated uncertainty are shown in shades of grey, by methodology, including Bayesian, Rogan-Gladen or Lang-Reiczigel. The top row shows 60% sensitivity, the middle row, 75%, and the bottom row 90%. Prior distribution assumptions are kept in line with simulation parameters. As test sensitivity increases the raw panel result more often is an overestimate. All three methods are able to correct the bias resulting from using the apparent prevalence as estimator for true prevalence.

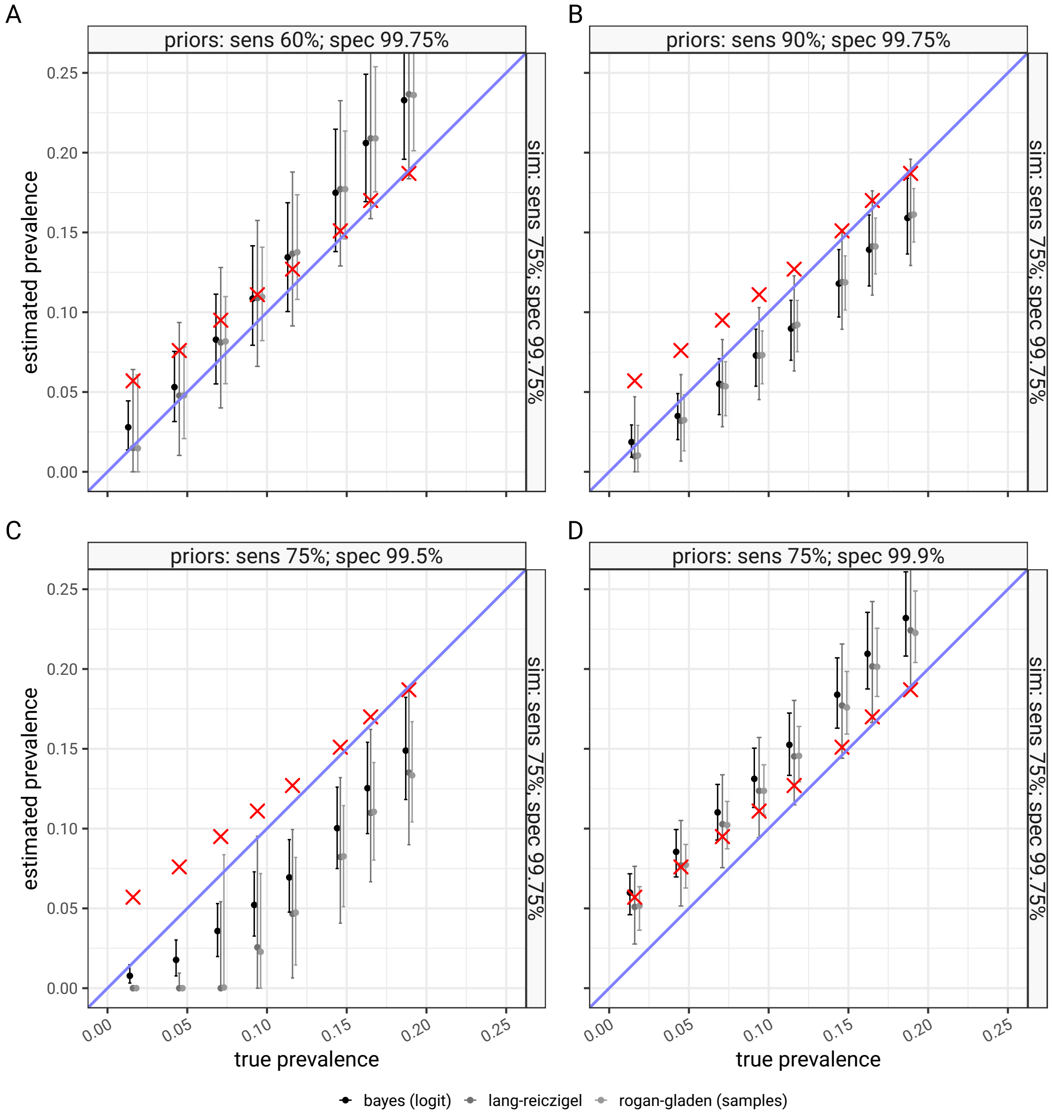


Figure 5: Adjustment of panel test error with extreme misspecification of test sensitivity and specificity parameters. The red crosses show apparent prevalence, and the estimates of adjusted prevalence with associated uncertainty are shown in shades of grey, by methodology, including Bayesian, Rogan-Gladen or Lang-Reiczigel. Subfigure A represents an overestimate of test sensitivity, B is an underestimate of test sensitivity, C is an underestimate of test specificity and D is an overestimate of specificity. The three adjustment methods fail in different ways, as informed priors are required to make accurate adjustments.

### 4 Discussion and Conclusions

We present three methods for correcting the bias in prevalence estimates in multiplex panel tests. These are implemented in an R package "testerror" [1]. These methods are all able to correct apparent prevalence estimates of components and panels with uncertain test sensitivity and specificity. No method is robust to misspecification, although Bayesian approaches can exclude some combinations of sensitivity and specificity based on the observed positive tests, (particularly combinations with comparatively low specificity, which would tend to make observations with low number of positives unlikely), the main adjustment requires accurate assessment of test parameters to produce an unbiased estimate but all methods are able to propagate imprecision in those estimates into the final prevalence estimates.

We have not formally quantified the performance of the various methods as there are a very large number of potential scenarios in which we could deploy them, and some of the methods are computationally expensive. On the evidence we have so far the three methods are each able to adjust the apparent prevalence of a simulated panel test results back to the true prevalence values the simulation is based on, with reasonable agreement between the methods. Within the software package released with this paper further validation of the methods is available.
